# Return Visits, Admission, and Mortality After Leaving Without Being Seen Among Emergency Department Patients With Behavioral Health Presentations

**DOI:** 10.64898/2026.10.14.26365554

**Authors:** Sarmed Ashoo, W. Frank Peacock, Christos Colovos, Jared C. Durieux

## Abstract

**Importance:** Patients with behavioral health presentations leave the emergency department (ED) before evaluation more often than other patients, but outcomes after leaving without being seen (LWBS) have not been described in this population.

**Objective:** To compare return visits, admission, and mortality after LWBS vs completed care among behavioral health ED episodes.

**Design, Setting, and Participants:** This retrospective episode-level cohort study used a multisystem deidentified electronic health record database of US health systems (data snapshot June 4, 2026). Participants were patients of all ages with an ED visit and same-day behavioral health diagnosis. Index visits (recorded January 26, 2001 to April 13, 2025) each opened a 30-day episode window; in-window were counted as outcomes, and patients could contribute episodes to either group. Data were analyzed in October 2026.

**Exposure:** LWBS (diagnosis code Z53.21 on a date with an ED encounter) vs completed care ending in discharge home.

**Main Outcomes and Measures:** The primary outcome was ED return visit within 7 days. Secondary outcomes were return visit within 3 and 30 days; admission on day 0, on days 1-7 (principal measure), and within 30 days; and all-cause death within 7 and 30 days, assessed among episodes from 2017 through 2023 using a verified death-record definition.

**Results:** The analysis included 1,305,812 episodes (12,248 LWBS; 1,293,564 completed care) from 525,490 patients; 666,930 episodes (51.1%) involved female patients and 66,733 (5.1%) involved patients younger than 18 years. Return visit within 7 days followed 24.5% of LWBS episodes vs 13.2% of completed-care episodes (risk ratio [RR], 1.86; 95% CI, 1.81-1.92), and admission on days 1-7 followed 17.1% vs 3.9% (RR, 4.41; 95% CI, 4.24-4.59). Among 2017-2023 episodes, 30-day mortality was 0.69% (70 of 10,141) after LWBS vs 0.32% (2,426 of 756,140) after completed care (RR, 2.15; 95% CI, 1.70-2.73). Results were similar under alternative episode definitions.

**Conclusions and Relevance:** Among patients with behavioral health ED presentations, LWBS was followed by higher rates of return visits, admission, and short-term mortality than completed care. These unadjusted findings describe outcome burden, not causal effects, and highlight patients who leave as a target for rapid follow-up.

**Key Points:** *Question:* Among patients with behavioral health emergency department presentations, are return visits, admission, and death more common after leaving without being seen than after completed care?

*Findings:* In this cohort study of 1,305,812 behavioral health emergency department episodes, 7-day return visits followed 24.5% of episodes in which the patient left without being seen vs 13.2% after completed care; admission on days 1-7 (17.1% vs 3.9%) and 30-day mortality (0.69% vs 0.32%) were also higher.

*Meaning:* Behavioral health patients who leave without being seen carry a greater short-term outcome burden, and the first days after departure are a practical window for outreach.

## Introduction

Mental health-related ED visits among US youth rose from 7.7% to 13.1% of pediatric ED visits (2011-2020), and mental, behavioral, and neurodevelopmental disorders account for nearly 6 million ED visits annually.^1,2^ Patients with behavioral health conditions wait longer, board longer, and leave emergency departments more often than other patients.^3,4^ The Centers for Medicare & Medicaid Services (CMS) tracks boarding for psychiatric admissions (OP-18c) but does not publish behavioral-health-specific rates for leaving without being seen, even as EDs care for a growing behavioral health population with limited inpatient capacity.^5^

Leaving without being seen occurs in approximately 1 to 3 of every 100 US ED visits and has increased since 2020.^6–9^ Because these patients leave before a clinician has evaluated them, a decision that might have led to admission or a safety plan is deferred to a later visit or never made. It is unknown how often these patients return, how often they are admitted, or whether short-term death differs from patients who complete ED care. Single-system studies report elevated return visit and admission rates after LWBS,^10–13^ and a population study from Ontario found higher 7-day death-or-hospitalization with longer ED waiting times,^14^ while more recent work found higher adverse-event risks after LWBS since 2020.^15^ A 2025 multicenter analysis found that 18.5% of LWBS visits were followed by a return visit within 7 days.^16^ However, none of these studies, nor a 2026 scoping review of 94 publications or a large pediatric cohort, isolated behavioral health presentations.^17,18^ Without such estimates, EDs cannot appropriately target rapid-return or outreach resources to these patients.

Using a multisystem electronic health record database with a large behavioral health population, we tested 3 episode-level hypotheses: that 7-day return visit and admission are higher after LWBS than after completed care, that a substantial share of admissions after LWBS occur on the same day, and that short-term mortality is higher after LWBS. We also report outcomes by sex, race, ethnicity, and age and describe the consistency of other premature-departure codes (absent without leave and against medical advice).

## Methods

### Study Design and Setting

We performed a retrospective cohort study using the Protege deidentified electronic health record database (data snapshot June 4, 2026), which contained records for 4.95 million patients from contributing US health systems. Before delivery data was de-identified through expert determination. An external review by Bluebonnet Ethical Review determined that this activity is appropriately classified as Not Human Subjects Research (NHSR) and that IRB review is not required. The protocol was finalized on October 6, 2026; the ED-encounter requirement, tie-date reclassification, and Pacific-state mortality correction were post hoc amendments. This study was supported through use of an internal Meta scaffold around Muse Wonder which assisted the authors in all the stages of the study. The study followed the Strengthening the Reporting of Observational Studies in Epidemiology (STROBE) and Reporting of Studies Conducted Using Observational Routinely-Collected Health Data (RECORD) reporting guidelines.^19,20^

### Selection of Participants

We included adult and pediatric behavioral health ED episodes, defined as a date with an ED encounter carrying a core behavioral health diagnosis (any *International Statistical Classification of Diseases, Tenth Revision, Clinical Modification* [ICD-10-CM] code F00-F99 or R45.850 , R45.851, or T14.91 on the same date. Each qualifying date was classified as LWBS (Z53.21) or completed care (ED encounter ending in discharge home) by that date’s own codes, and LWBS required an ED encounter on that date. Episodes were built per patient in chronological order. A visit began a new 30-day episode (washout) window only if it fell outside any earlier window; a visit inside an open window was counted as an outcome of that episode and never began its own. Cohort assembly is shown in Figure 1. Sensitivity analyses used 7-day washout and every-visit counting with no washout (eMethods in Supplement 1).

**Figure 1.**
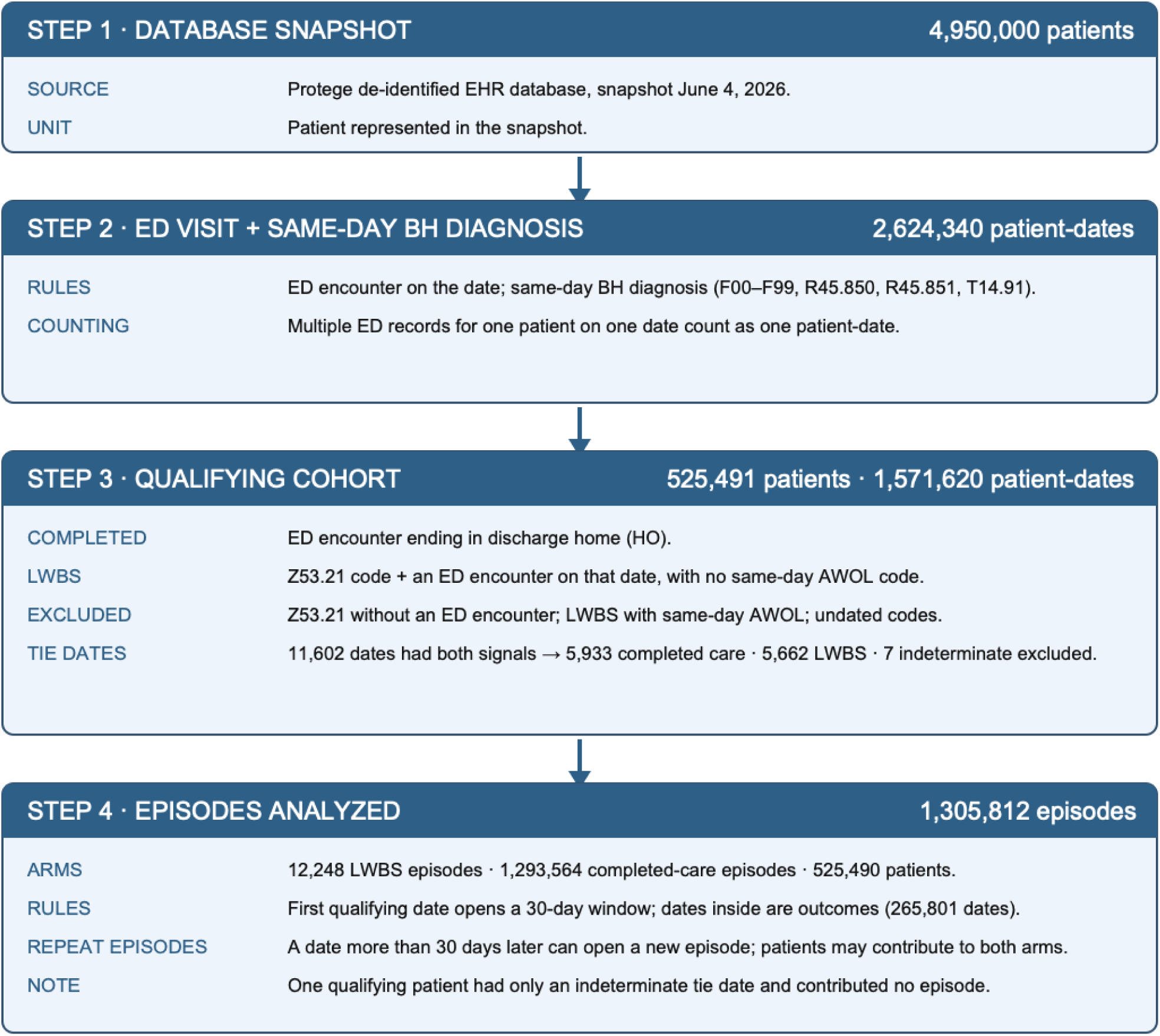
Flow Diagram of Episode Cohort Assembly (30-Day Episode Window). Each step shows the unit and count after that step’s rules are applied. Steps 1 and 3 count patients, Step 2 counts patient-dates, and Step 4 counts date-defined episodes. One qualifying patient had only an indeterminate tie date and contributed no episode, so the episode analysis includes 525,490 of the 525,491 qualifying patients. BH indicates behavioral health; ED, emergency department; LWBS, left without being seen.

Same-day tie dates (11,602) had both an LWBS code and a completed-care encounter. Because diagnoses were recorded by patient-date, not encounter, all ED encounters on the date were assessed and the date was classified once. Dates accounted for by discharge-home or inpatient encounters were classified as completed care (5,933); dates with a separate explicit premature-departure encounter or an additional unaccounted encounter were classified as LWBS (5,662); and 7 dates without evaluation evidence were excluded. Premature-departure codes were used only to adjudicate these Z53.21 dates, not as independent LWBS criteria. The complete algorithm is provided in Supplement 1.

The 30-day episode window was the primary comparison because it prevents overlapping follow-up and prevents any visit from being both an outcome and the start of another episode. The every-visit calculation was performed separately to match the visit-counting approach of Evans et al^16^ for descriptive comparison.

Absent-without-leave and against-medical-advice codes were examined by case review but not analyzed as separate groups because the codes were used inconsistently.

### Measurements

Sex, race, ethnicity, and age band were recorded as stratification variables. Age was calculated at each episode and banded as younger than 18 years, 18–29, 30–39, 40–49, 50–59, 60–69, and 70 years or older, so a patient could contribute episodes to different age bands over time. Sex, race, and ethnicity came from the electronic health record as recorded by each contributing health system; race and ethnicity were examined because differences in ED waiting and departure before evaluation by race and ethnicity have been reported.^6,8^ When a patient’s demographic records disagreed, specific values were preferred over “Other” or blank entries. Episodes whose patients still had conflicting values were retained in the overall analyses but not included in that variable’s labeled strata. The episode counts omitted from each labeled set of strata are reported in the Table 1 footnote. Counts below 11 are not reported, with additional complementary suppression to prevent back-calculation.

**Table 1.** Characteristics of Behavioral Health Emergency Department Episodes by Disposition.

| Characteristic | LWBS episodes<br>(n = 12,248) | Completed-care episodes<br>(n = 1,293,564) |
| --- | --- | --- |
| Episodes, No. | 12,248 | 1,293,564 |
| Patients, No.<sup>a</sup> | 11,514 | 522,130 |
| Female sex, No. (%) | 5,954 (48.6) | 660,976 (51.1) |
| Male sex, No. (%) | 6,262 (51.1) | 628,750 (48.6) |
| White race, No. (%) | 8,040 (65.6) | 818,586 (63.3) |
| Black race, No. (%) | 2,838 (23.2) | 343,958 (26.6) |
| Asian race, No. (%) | 208 (1.7) | 20,840 (1.6) |
| Other race, No. (%) | 805 (6.6) | 83,813 (6.5) |
| Non-Hispanic ethnicity, No. (%) | 10,339 (84.4) | 1,106,386 (85.5) |
| Hispanic ethnicity, No. (%) | 677 (5.5) | 69,301 (5.4) |
| Other ethnicity, No. (%) | 739 (6.0) | 83,935 (6.5) |
| Age <18 y, No. (%) | 279 (2.3) | 66,454 (5.1) |
| Age 18-29 y, No. (%) | 2,511 (20.5) | 307,696 (23.8) |
| Age 30-39 y, No. (%) | 3,027 (24.7) | 298,213 (23.1) |
| Age 40-49 y, No. (%) | 2,292 (18.7) | 225,966 (17.5) |
| Age 50-59 y, No. (%) | 1,828 (14.9) | 190,562 (14.7) |
| Age 60-69 y, No. (%) | 1,311 (10.7) | 114,560 (8.9) |
| Age ≥70 y, No. (%) | 999 (8.2) | 90,072 (7.0) |

Percentages are episode-weighted: each episode carries the characteristics of the patient who had that episode, so patients contributing multiple episodes are represented multiple times. Episodes not shown in labeled strata because the patient had conflicting or missing values: sex, 32 left without being seen (LWBS) and 3,838 completed-care episodes; race, 357 and 26,367; ethnicity, 493 and 33,942. One LWBS episode and 41 completed-care episodes had unknown age band.
<sup>a</sup>Patients could contribute episodes to both groups. Of the 525,490 patients, 513,976 (97.8%) contributed only completed-care episodes, 3,360 (0.6%) only LWBS episodes, and 8,154 (1.6%) both; patients in both groups contributed 8,842 LWBS and 51,460 completed-care episodes.

### Outcomes

The primary outcome was ED return visit within 7 days after the episode date. Secondary outcomes were return visit within 3 calendar days (days 1–3 after the episode date; the data contain dates only, with no visit timestamps, so returns are counted by calendar date rather than elapsed time) and within 30 days; inpatient admission on day 0, on days 1–7, pooled over days 0–7, and within 30 days; and all-cause death within 7 and 30 days. Day-0 admissions are reported as same-day co-occurrence and shown separately because, with date-only data, a same-day admission cannot be distinguished from continuation of the index encounter or from a later same-day return after departure. The days 1–7 contrast is the cleaner measure of admission after the index date. A patient could have admissions in both the day-0 and days 1–7 windows, so those windows are not mutually exclusive and do not sum to the pooled 7-day rate.

All-cause death within 7 and 30 days was counted under a prespecified verified definition. A death counted only if it appeared in the death-record file with a death date of January 1, 2017, or later and with supporting evidence from Social Security verification, a confidence score of 94 or higher, or, outside Washington, Alaska, Montana, Idaho, Hawaii, and Oregon, a matching death date in the patient’s demographic record within 31 days; patients from those states were not excluded. Patient state was assigned as the patient’s most frequently recorded state because patients could have more than one state recorded and no validated latest-record rule exists. Deaths followed by an emergency department visit, inpatient stay, procedure, or surgery more than 180 days after the recorded death date were excluded. The mortality analysis was restricted to episodes from January 1, 2017, through December 31, 2023, so that the full 30-day follow-up window fell within the period of usable death capture; death capture increased sharply in 2017 and the last captured death was September 21, 2024. Pediatric return visit and admission were prespecified subgroup outcomes. Pediatric deaths are not reported because counts were below the reporting threshold.

### Statistical Analysis

Return visit, admission, and mortality proportions were episode-level rates calculated directly, with Wilson score 95% CIs.^21^ We report unadjusted risk differences with Newcombe hybrid score 95% CIs and risk ratios with log-based 95% CIs.^22^ Contrasts are unadjusted comparisons of outcomes following LWBS versus completed-care episodes. They do not establish causal effects of departure. Episodes from the same patient are not statistically independent. The main intervals treat episodes as independent. Cluster-robust analyses clustered by patient produced materially the same estimates. Subgroup rates by sex, race, ethnicity, and age band are reported descriptively (eTable 1 in Supplement 1). Interaction analyses used Poisson likelihood-ratio homogeneity tests, with Bonferroni correction across 24 tests. Sensitivity analyses excluded patients whose only death signal was a death date in the patient’s demographic record without an entry in the death-record file; mortality results were unchanged. Patients with no records after an episode were counted as having no event, which cannot distinguish a true absence of events from care outside contributing systems. Deaths remain in the denominators of return visit and admission rates and are reported as separate endpoints. Site-level clustering could not be addressed because no site identifier was available for index encounters. Analyses were performed using Presto version 0.300 and Python version 3 (Python Software Foundation).

## Results

### Patient Characteristics

The primary analysis included 12,248 LWBS and 1,293,564 completed-care episodes from 525,490 patients (Figure 1). Of these patients, 240,746 (45.8%) contributed more than 1 episode, and 8,154 (1.6%) contributed episodes to both groups (Table 1). Demographic percentages are episode-weighted. Female patients accounted for 48.6% of LWBS episodes vs 51.1% of completed-care episodes. White patients accounted for 65.6% of LWBS episodes vs 63.3% of completed-care episodes, Black patients 23.2% vs 26.6%, and non-Hispanic patients 84.4% vs 85.5% (Table 1). By age band, LWBS episodes were 2.3% pediatric (<18 years), 20.5% aged 18-29, 24.7% aged 30-39, and 8.2% aged 70 years or older, vs 5.1%, 23.8%, 23.1%, and 7.0% for completed care (eFigure 1 in Supplement 1).

### Return Visits and Admission

At 7 days, 3,004 of 12,248 LWBS episodes (24.5%) were followed by an ED return vs 170,129 of 1,293,564 completed-care episodes (13.2%; risk difference, 11.4 [95% CI, 10.6-12.1] percentage points; risk ratio, 1.86 [95% CI, 1.81-1.92]; Table 2 and Figure 2). The gap was present within 3 days (19.4% vs 9.0%) and at 30 days (36.8% vs 25.1%). Admission on days 1-7 occurred after 2,092 LWBS episodes (17.1%) vs 50,123 completed-care episodes (3.9%; risk ratio, 4.41 [95% CI, 4.24-4.59]). Same-day (day 0) admission co-occurrence was 18.9% vs 1.6% and pooled 7-day admission 26.0% vs 4.8%. At 30 days, admission was 31.1% vs 8.9%. With 7-day washout, 7-day return was 26.7% vs 14.6% (risk ratio, 1.84); with every-visit counting, 30.5% vs 16.6% (risk ratio, 1.84).

**Figure 2.**
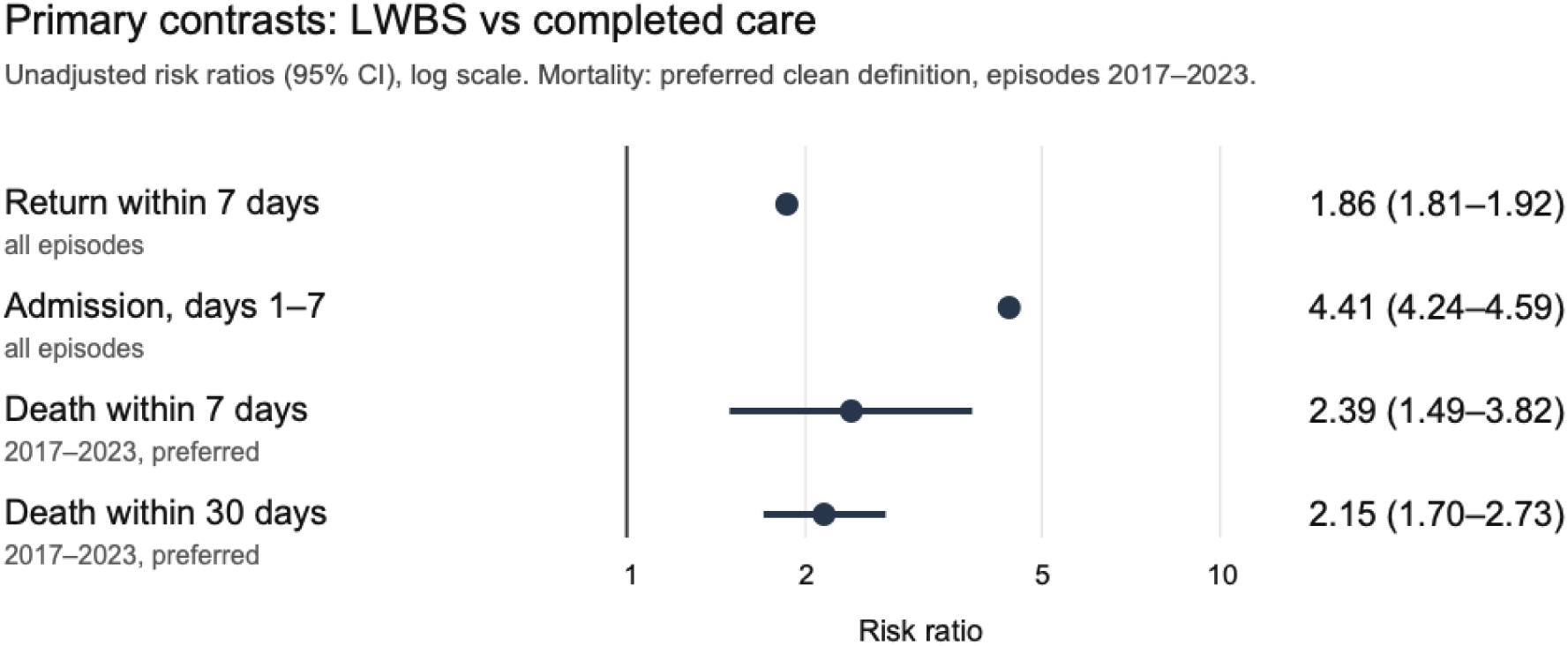
Risk Ratios for Return Visit, Admission, and Mortality After LWBS vs Completed Care Episodes. Return and admission use all episodes under the 30-day window; mortality uses the preferred Pacific-corrected analysis among episodes from 2017 through 2023. Points show log-based risk ratios with 95% CIs. LWBS indicates left without being seen.

**Table 2.** Return Visit, Admission, and Mortality After LWBS vs Completed Care, Episode Cohort.

| Outcome | LWBS<br>(n = 12,248) | Completed care<br>(n = 1,293,564) | Risk difference,<br>percentage<br>points (95% CI) | Risk ratio<br>(95% CI) |
| --- | --- | --- | --- | --- |
| Return visit, 3 d | 2,377; 19.4%<br>(18.7-20.1) | 116,182; 9.0%<br>(8.9-9.0) | 10.4<br>(9.7 to 11.1) | 2.16<br>(2.08-2.24) |
| Return visit, 7 d (primary) | 3,004; 24.5%<br>(23.8-25.3) | 170,129; 13.2%<br>(13.1-13.2) | 11.4<br>(10.6 to 12.1) | 1.86<br>(1.81-1.92) |
| Return visit, 30 d | 4,511; 36.8%<br>(36.0-37.7) | 324,086; 25.1%<br>(25.0-25.1) | 11.8<br>(10.9 to 12.6) | 1.47<br>(1.44-1.50) |
| Admission, day 0<br>(same-day co-occurrence) | 2,314; 18.9%<br>(18.2-19.6) | 20,581; 1.6%<br>(1.6-1.6) | 17.3<br>(16.6 to 18.0) | 11.87<br>(11.42-12.35) |
| Admission, days 1-7 | 2,092; 17.1%<br>(16.4-17.8) | 50,123; 3.9%<br>(3.8-3.9) | 13.2<br>(12.5 to 13.9) | 4.41<br>(4.24-4.59) |
| Admission, 7 d (pooled) | 3,189; 26.0%<br>(25.3-26.8) | 62,700; 4.8%<br>(4.8-4.9) | 21.2<br>(20.4 to 22.0) | 5.37<br>(5.21-5.54) |
| Admission, 30 d | 3,814; 31.1%<br>(30.3-32.0) | 115,212; 8.9%<br>(8.9-9.0) | 22.2<br>(21.4 to 23.1) | 3.50<br>(3.40-3.59) |
| Death, 7 d<sup>a</sup> | 18; 0.18%<br>(0.11-0.28) | 562; 0.07%<br>(0.07-0.08) | 0.10<br>(0.04 to 0.21) | 2.39<br>(1.49-3.82) |
| Death, 30 d<sup>a</sup> | 70; 0.69%<br>(0.55-0.87) | 2,426; 0.32%<br>(0.31-0.33) | 0.37<br>(0.23 to 0.55) | 2.15<br>(1.70-2.73) |

Left without being seen (LWBS) and completed care are shown as number of events, percentage, and Wilson score 95% CI. Risk differences use Newcombe hybrid score 95% CIs; risk ratios use log-based 95% CIs. Day-0 and days 1–7 admission windows are not mutually exclusive. Return and admission rows use all episodes.
<sup>a</sup>Mortality rows use the verified death-record definition with Pacific-state correction (see Methods) among episodes from 2017 through 2023 (LWBS, n = 10,141; completed care, n = 756,140), not the all-episode denominators in the column headers.

### Mortality

Among episodes from 2017 through 2023, 30-day mortality followed 70 of 10,141 LWBS episodes (0.69%) vs 2,426 of 756,140 completed-care episodes (0.32%; risk difference, 0.37 [95% CI, 0.23 to 0.55] percentage points; risk ratio, 2.15 [95% CI, 1.70-2.73]; Table 2). Seven-day mortality was 18 of 10,141 LWBS episodes (0.18%) vs 562 of 756,140 completed-care episodes (0.07%; risk ratio, 2.39 [95% CI, 1.49-3.82]). Removing patient-record confirmation in Washington, Alaska, Montana, Idaho, Hawaii, and Oregon removed 54 completed-care death outcomes and no LWBS death outcomes in this window. Mortality analyses were descriptive and were not powered for formal comparison.

### Pediatric Subgroup

Among episodes for patients younger than 18 years in the 2017–2023 subgroup analysis, 7-day return occurred after 28 of 234 LWBS episodes (12.0%) vs 3,215 of 37,746 completed-care episodes (8.5%), and admission on days 1-7 after 23 (9.8%) vs 1,259 (3.3%) (eTable 1 and eFigure 2 in Supplement 1). The pattern was similar to that in adults at lower absolute levels. Pediatric mortality counts were suppressed under the small-cell policy, so no pediatric mortality contrast is estimated.

### Validation and Sensitivity Analyses

The counting rule changed the denominators but not the qualitative findings (Table 3). Seven-day return risk ratios were nearly identical under the 30-day episode window (1.86), the 7-day episode window (1.84), and every-visit counting (1.84). Admission on days 1-7 was somewhat stronger with the 30-day window (risk ratio, 4.41) than with the 7-day window (4.13) or every-visit counting (3.81). In 2017-2023, the return association differed by race, and the admission association differed by race and ethnicity, after Bonferroni correction. No other heterogeneity was detected (eTable 2 in Supplement 1).

**Table 3.** Main Contrasts Under 3 Counting Rules.

| Counting rule | LWBS | Completed care | Return within 7 d, RR (95% CI) | Admission days 1-7, RR (95% CI) | Death within 30 d, RR (95% CI) |
| --- | --- | --- | --- | --- | --- |
| 30-day episode window (primary) | 12,248 episodes | 1,293,564 episodes | 1.86 (1.81-1.92) | 4.41 (4.24-4.59) | 2.15 (1.70-2.73)<sup>a</sup> |
| 7-day episode window | 13,678 episodes | 1,429,182 episodes | 1.84 (1.79-1.89) | 4.13 (3.98-4.30) | Not estimated |
| Every visit counted (no episode window) | 15,505 visits | 1,556,108 visits | 1.84 (1.79-1.88) | 3.81 (3.67-3.95) | Not estimated |

Counts are episodes for the 30-day and 7-day rows and visits for the every-visit row. Risk ratios (RR) use log-based 95% CIs. The 30-day episode-window row is the primary comparison; the every-visit row follows the visit-counting approach of Evans et al for the left without being seen (LWBS benchmark).
<sup>a</sup>Mortality is the 30-day estimate with Pacific-state correction among episodes from 2017 through 2023; it was not re-estimated under the other counting rules.

## Discussion

In this multisystem cohort of 1,305,812 behavioral health ED episodes, leaving without being seen was followed by return within 7 days in 24.5% vs 13.2% after completed care and by admission on days 1-7 in 17.1% vs 3.9%, with the same direction under 7-day washout and every-visit counting. Reclassifying same-day tie dates by encounter evidence strengthened the admission contrast. Because a same-day admission cannot be distinguished from continuation of the index encounter in date-only data, the days 1-7 contrast (risk ratio, 4.41) carries the admission finding. This pattern is consistent with prior reports of early return after LWBS^10–13,23^ and identifies the first days after departure as a practical window for outreach, crisis-line linkage, or return precautions tailored to behavioral health presentations, approaches that have reduced suicidal behavior after ED care.^24,25^

Evans et al^16^ studied LWBS among adults using Epic Cosmos data from 2023. Of 345,938 LWBS visits, 18.5% were followed by an ED return within 7 days and 28.4% within 30 days, with a median time to return of 46.6 hours. That study included no completed-care comparator, no episode construction, and no mortality outcome, so it could not show how much more often patients return than similar patients who complete care, or whether short-term death differs.

In the closest like-for-like count (every-visit counting with no washout), 30.5% of behavioral health LWBS visits (4,732 of 15,505) were followed by a return within 7 days and 43.8% within 30 days, above the 18.5% and 28.4% reported by Evans et al. These levels should not be read as a head-to-head comparison: the populations (behavioral health, all ages vs general adults), data sources, calendar periods, and LWBS recording practices differ. The comparison does suggest that return after behavioral health LWBS is at least as frequent as after general-population LWBS and that the excess over completed care persists when repeat visits are handled explicitly. The shared early-return timing (median under 2 days in Evans et al; 19.4% by day 3 here) points to the same window for outreach.

Short-term mortality was higher after LWBS episodes among episodes from 2017 through 2023 (30-day risk ratio, 2.15 [95% CI, 1.70-2.73]). Absolute risk remained small. Mortality after ED presentation for self-harm and suicidal ideation is known to be elevated,^26^ and whether deaths after behavioral health LWBS reflect suicide, overdose, or medical causes could not be determined because cause of death was not available.

Age distributions were broadly similar between groups, so the outcome differences are unlikely to be explained by a markedly different age composition, although children formed a smaller share of LWBS episodes. Interaction testing found heterogeneity by race for return visits and by race and ethnicity for admission, but no detectable heterogeneity for mortality or for the other return and admission dimensions examined. Children showed the same pattern as adults at lower absolute levels (7-day return, 12.0% vs 8.5%; admission on days 1-7, 9.8% vs 3.3%). Pediatric mortality contrast was not estimated in this study.

Finally, absent-without-leave and against-medical-advice codes did not identify consistent populations in this database. Absent-without-leave codes mixed records linked to admissions, walkouts, and records without encounter evidence. Against-medical-advice codes mixed signed refusals, elopements, and LWBS-like departures. Patients who leave against medical advice are a heterogeneous group in other ED settings as well.^27^ This heterogeneity, established by case review before outcome analysis, has implications for multisystem ED research that relies on these codes.

### Limitations

Our study has several limitations. First, patients who leave differ from patients who complete care in acuity, substance use, housing, and other unmeasured factors. They do not establish causal effects of departure, and the direction of selection bias is unknown. Second, patients with fewer clinical notes are underrepresented, so the LWBS group overrepresents patients with more frequent contact. This could bias rates in either direction. Third, same-day admissions may continue the index episode rather than represent new outcomes. We therefore report day 0 separately and base the admission finding on days 1–7. Fourth, mortality may still be undercounted even after restricting the analysis to 2017-2023 and removing unreliable Pacific-state confirmations. Deaths recorded only in the patient’s demographic record were not counted, and recorded death dates may lag the true date. The effect on the risk ratio is uncertain. Fifth, episodes from the same patient are correlated. Cluster-robust analyses were materially similar, but unmeasured patient-level differences remain. Sixth, observation stays are captured where known. Short-stay admissions managed as observation may be missed asymmetrically, with uncertain observation capture after LWBS. Seventh, absolute rates are not national estimates. The database is enriched for behavioral health and geographically concentrated, and payer information was not available, so insurance status could not be examined. Eighth, same-day behavioral health coding is a documentation behavior and may be less complete for visits ending before evaluation. Finally, pediatric mortality is too rare to contrast under the small-cell policy.

Several of these limitations could be addressed by prospective capture of departure circumstances and waiting times, linkage to enrollment data for out-of-system outcomes, and evaluation of systematic, patient-centered rapid-return contact in the days after LWBS.

### Conclusions

In this large multisystem cohort of behavioral health ED episodes, leaving without being seen was followed by substantially higher rates of return visits, admission, and short-term mortality than completed ED care, and children showed a similar pattern for return visits and admission.

Findings were robust to alternative episode definitions and to encounter-level reclassification of same-day tie dates. Prospective studies should assess whether rapid-return pathways and next-day outreach after behavioral health LWBS reduce return visits, admission, and mortality.

## Data Availability

The Protege data are not publicly available and access is governed by the data owner. The study protocol,
statistical analysis plan, code lists and aggregate results are available from the Corresponding author on
reasonable request..

## Author Contributions

Dr Ashoo had full access to all of the data in the study. Concept and design: Ashoo. Acquisition, analysis, or interpretation of data: Ashoo, Peacock. Drafting of the manuscript: Ashoo. Critical review of the manuscript for important intellectual content: Ashoo, Peacock, Colovos. Statistical analysis: Ashoo, Durieux.

## Funding/Support

This work was supported by Meta Superintelligence Labs

## Conflict of Interest Disclosures

All authors declare the following competing interests: this work was completed by the authors while employed at, or while collaborating with, Meta.

## Prior Presentations

None

## Supplement 1

### eMethods

#### Tie-Date Classification

When a date contained both an LWBS diagnosis code and an ED visit ending in discharge home, the date was classified once because the diagnosis code could not be linked to an individual ED visit. The ED visits recorded on that date were reviewed in the following order: (1) a single ED visit classified the date as completed care; (2) a visit ending because the patient left against medical advice or without leave classified the date as LWBS; (3) a date on which every ED visit ended in discharge home was classified as completed care; (4) a date fully accounted for by ED visits ending in discharge home or inpatient admission, with no visit disposition missing or recorded as another status, was classified as completed care; (5) a date with no same-day clinical note, laboratory result, or procedure record was excluded; and (6) any remaining visit with a missing or other recorded disposition classified the date as LWBS. These departure categories were used only to resolve dates with the LWBS diagnosis code; they did not define LWBS on their own.

#### Unaccounted-Encounter Sensitivity Analysis

The 3,003 dates classified as LWBS because of an additional ED visit with no recorded disposition or a disposition recorded as another status were instead excluded as indeterminate, and episodes were reconstructed. This yielded 9,813 LWBS and 1,293,921 completed-care episodes among 524,963 patients. Return within 7 days occurred after 25.3% of LWBS episodes vs 13.2% of completed-care episodes (risk ratio, 1.92), admission on days 1-7 after 18.2% vs 3.9% (risk ratio, 4.69), and, among episodes from 2017-2023, death within 30 days after 0.71% vs 0.32% (risk ratio, 2.21), compared with risk ratios of 1.86, 4.41, and 2.15, respectively, in the primary analysis. The subgroup conclusions were unchanged: the return association differed by race, the admission association differed by race and ethnicity, and no other differences were detected.

#### Repeated Episodes From the Same Patient

Because some patients contributed more than 1 episode, including 8,154 patients who contributed episodes to both groups, we repeated the analysis while accounting for repeated episodes from the same patient. Risk differences and risk ratios for all outcomes in Table 2 were materially unchanged.

#### Every-Visit Sensitivity Analysis

Each qualifying LWBS or completed-care visit was counted as an index visit without grouping visits into episodes, following the approach of Evans et al. Counts below the reporting threshold were suppressed, with additional suppression where needed to prevent their calculation from other reported values.

---

**eTable 1.** Subgroup Outcome Rates, Episode-Weighted (Episodes 2017–2023, Pacific-Corrected Mortality)

| Stratum | LWBS return<br>7d | Completed<br>return 7d | LWBS<br>admission<br>d1-7 | Completed<br>admission<br>d1-7 | LWBS death<br>30d | Completed<br>death 30d |
| --- | --- | --- | --- | --- | --- | --- |
| Female | 1,073/4,925<br>(21.8%) | 46,487/370,616<br>(12.5%) | 804/4,925<br>(16.3%) | 16,101/370,616<br>(4.3%) | 36/4,925<br>(0.7%) | 1,078/370,616<br>(0.3%) |
| Male | 1,401/5,191<br>(27.0%) | 56,877/383,821<br>(14.8%) | 951/5,191<br>(18.3%) | 17,190/383,821<br>(4.5%) | 34/5,191<br>(0.7%) | 1,348/383,821<br>(0.4%) |
| White | 1,545/6,729<br>(23.0%) | 64,727/486,834<br>(13.3%) | 1,209/6,729<br>(18.0%) | 23,796/486,834<br>(4.9%) | 55/6,729<br>(0.8%) | 1,940/486,834<br>(0.4%) |
| Black | 660/2,265<br>(29.1%) | 26,495/185,612<br>(14.3%) | 348/2,265<br>(15.4%) | 6,754/185,612<br>(3.6%) | Suppressed | 386/185,612<br>(0.2%) |
| Asian | 39/165 (23.6%) | 1,772/13,489<br>(13.1%) | 35/165 (21.2%) | 539/13,489<br>(4.0%) | Suppressed | 27/13,489<br>(0.2%) |
| Other race | 136/682<br>(19.9%) | 7,380/54,120<br>(13.6%) | 124/682<br>(18.2%) | 1,735/54,120<br>(3.2%) | Suppressed | 62/54,120<br>(0.1%) |
| Non-Hispanic | 2,118/8,516<br>(24.9%) | 85,136/619,376<br>(13.7%) | 1,484/8,516<br>(17.4%) | 28,960/619,376<br>(4.7%) | 63/8,516<br>(0.7%) | 2,290/619,376<br>(0.4%) |
| Hispanic | 121/582<br>(20.8%) | 6,440/45,604<br>(14.1%) | 94/582 (16.2%) | 1,501/45,604<br>(3.3%) | 0/582 (0.0%) | 22/45,604<br>(0.0%) |
| Other/unknown<br>ethnicity | 115/611<br>(18.8%) | 7,831/68,226<br>(11.5%) | 113/611<br>(18.5%) | 2,202/68,226<br>(3.2%) | Suppressed | 93/68,226<br>(0.1%) |
| <18 y | 28/234 (12.0%) | 3,215/37,746<br>(8.5%) | 23/234 (9.8%) | 1,259/37,746<br>(3.3%) | 0/234 (0.0%) | Suppressed |
| 18-29 y | 439/2,042<br>(21.5%) | 20,711/167,110<br>(12.4%) | 252/2,042<br>(12.3%) | 5,319/167,110<br>(3.2%) | Suppressed | 62/167,110<br>(0.0%) |
| 30-39 y | 600/2,562<br>(23.4%) | 25,034/178,190<br>(14.0%) | 345/2,562<br>(13.5%) | 6,197/178,190<br>(3.5%) | Suppressed | 108/178,190<br>(0.1%) |
| 40-49 y | 516/1,848<br>(27.9%) | 18,357/129,599<br>(14.2%) | 312/1,848<br>(16.9%) | 5,008/129,599<br>(3.9%) | Suppressed | 139/129,599<br>(0.1%) |
| 50-59 y | 408/1,520<br>(26.8%) | 15,785/108,609<br>(14.5%) | 298/1,520<br>(19.6%) | 5,183/108,609<br>(4.8%) | Suppressed | 262/108,609<br>(0.2%) |
| 60-69 y | 285/1,097<br>(26.0%) | 11,442/73,153<br>(15.6%) | 266/1,097<br>(24.2%) | 4,794/73,153<br>(6.6%) | 12/1,097<br>(1.1%) | 411/73,153<br>(0.6%) |
| 70+ y | 206/837<br>(24.6%) | 9,103/61,712<br>(14.8%) | 265/837<br>(31.7%) | 5,621/61,712<br>(9.1%) | 36/837 (4.3%) | 1,439/61,712<br>(2.3%) |

Cells show events/episodes (percentage); percentages were calculated directly from the counts shown. Cells labeled “Suppressed” had death counts below the reporting threshold (fewer than 11); complementary suppression was applied to prevent back-calculation. LWBS indicates left without being seen.

**eTable 2.** Episode-Level Interaction (Effect-Modification) Summary After Bonferroni Correction (Episodes 2017–2023)

| Dimension | Episode: return | Episode: admission | Episode: mortality |
| --- | --- | --- | --- |
| Sex | No difference detected<br>($\chi^2=1.34$, df=1; P=.248;<br>adjusted P=1.00) | No difference detected<br>($\chi^2=2.99$, df=1; P=.084;<br>adjusted P=1.00) | No difference detected<br>($\chi^2=1.51$, df=1; P=.219;<br>adjusted P=1.00) |
| Race | Differs<br>($\chi^2=18.45$, df=3; P<.001;<br>adjusted P=.009) | Differs<br>($\chi^2=23.59$, df=3; P<.001;<br>adjusted P<.001) | No difference detected<br>($\chi^2=1.10$, df=3; P=.778;<br>adjusted P=1.00) |
| Ethnicity | No difference detected<br>($\chi^2=5.89$, df=2; P=.053;<br>adjusted P=1.00) | Differs<br>($\chi^2=21.43$, df=2; P<.001;<br>adjusted P<.001) | No difference detected<br>($\chi^2=9.90$, df=2; P=.007;<br>adjusted P=.170) |
| Age band | No difference detected<br>($\chi^2=12.04$, df=6; P=.061;<br>adjusted P=1.00) | No difference detected<br>($\chi^2=10.52$, df=6; P=.105;<br>adjusted P=1.00) | No difference detected<br>($\chi^2=5.16$, df=6; P=.523;<br>adjusted P=1.00) |

Poisson likelihood-ratio tests compared a null model with a common LWBS-vs-completed-care risk ratio across strata, fitted by maximum likelihood, with an alternative model allowing stratum-specific risk ratios. P values use the chi-square distribution with degrees of freedom equal to the number of strata minus 1 and were Bonferroni-adjusted across 24 outcome–dimension tests. “Differs” means adjusted P < .05; “No difference detected” means adjusted P ≥ .05 and does not prove that the associations are equal. LWBS indicates left without being seen.

**eFigure 1.**
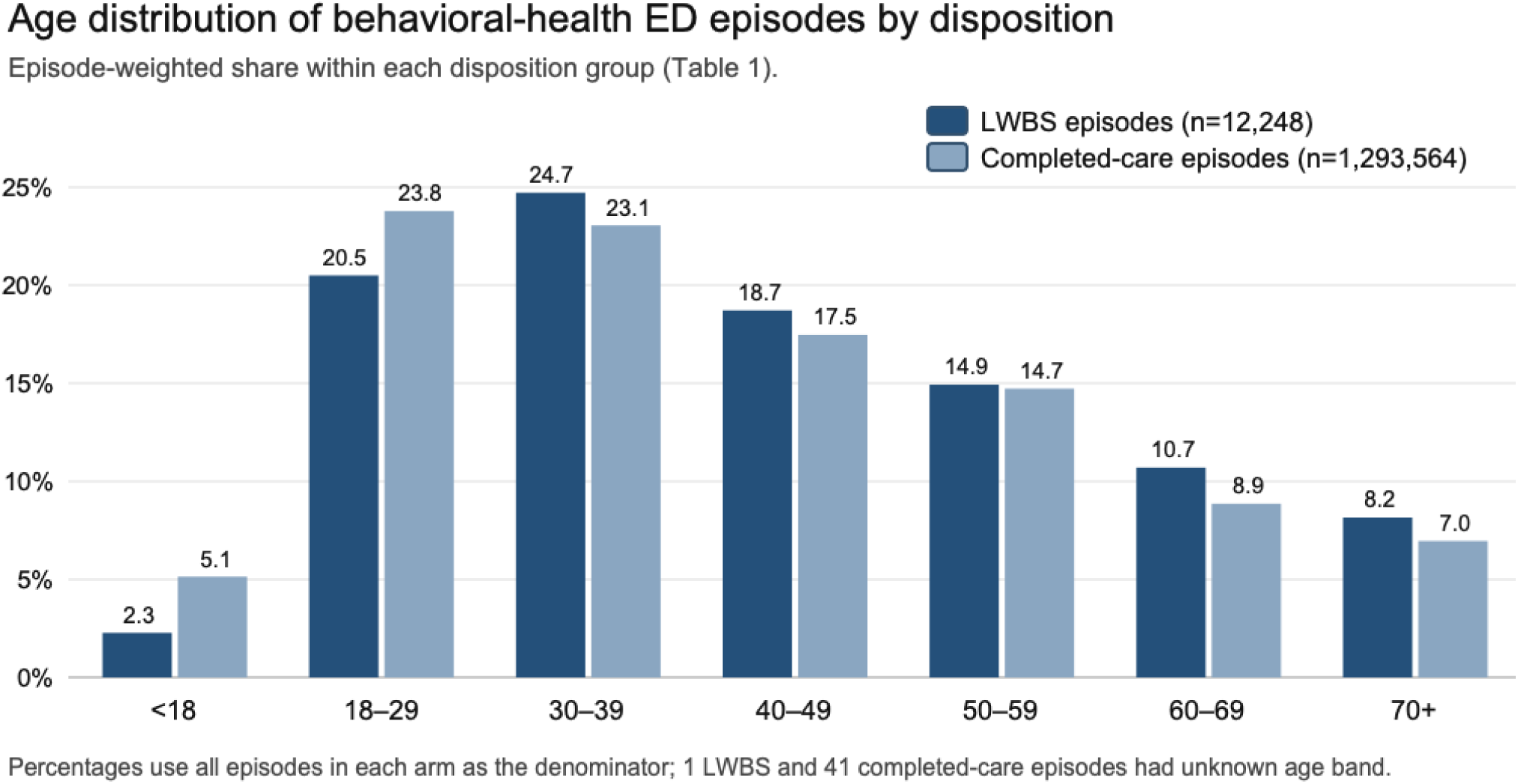
Age Distribution of Behavioral Health ED Episodes by Disposition. Grouped bars show the episode-weighted share of LWBS and completed-care episodes in each age band (Table 1). Ages 30–39 years form the largest group among LWBS episodes and ages 18–29 years among completed-care episodes; pediatric episodes form a smaller share of LWBS than completed-care episodes. LWBS indicates left without being seen.

**eFigure 2.**
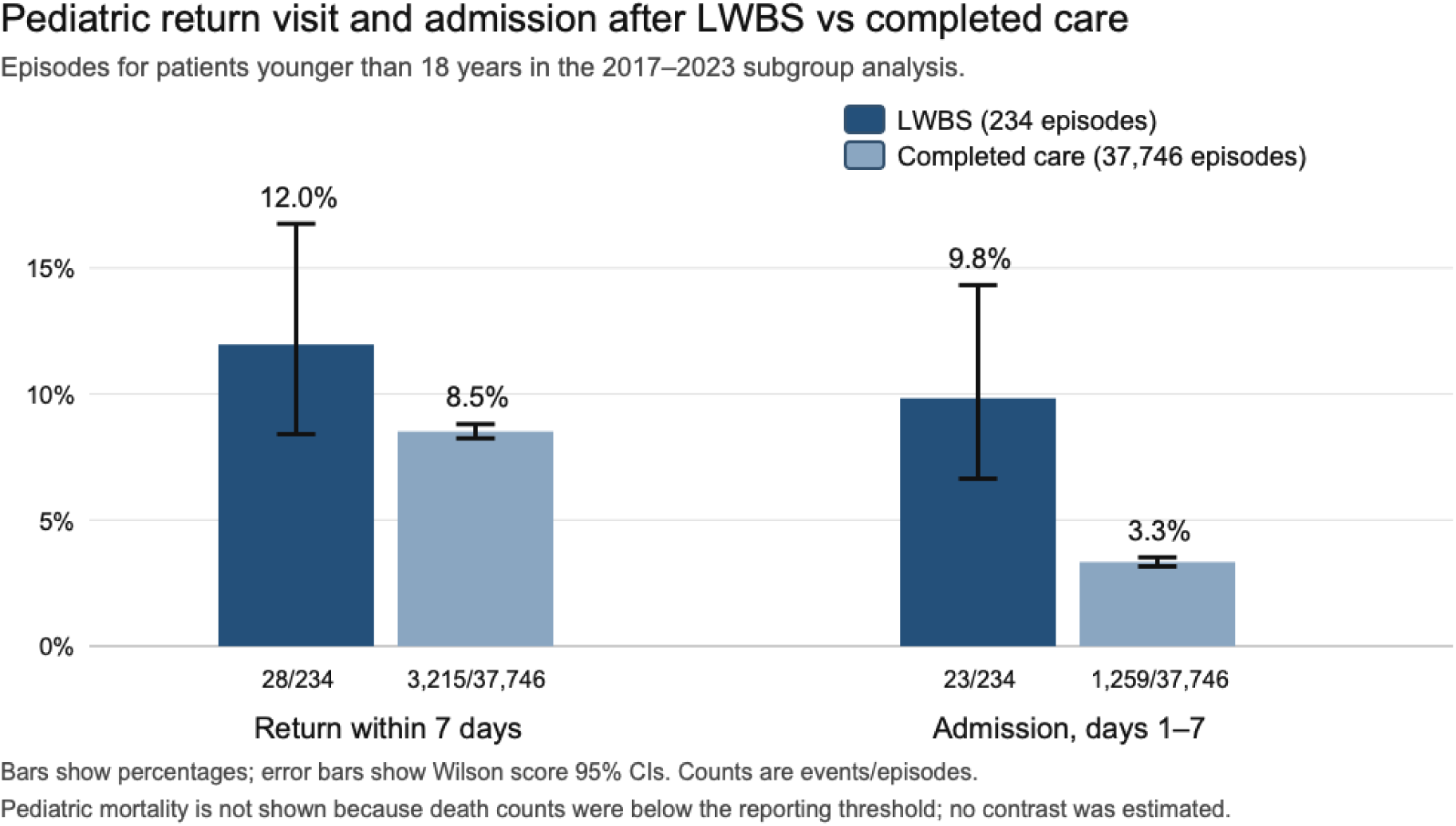
Pediatric Return Visit and Admission After LWBS vs Completed Care. Among pediatric episodes in the 2017–2023 subgroup analysis, return within 7 days and admission on days 1–7 are shown for LWBS and completed care. Bars show percentages; error bars show Wilson score 95% CIs. Pediatric mortality is not shown because death counts were below the reporting threshold and no contrast was estimated. LWBS indicates left without being seen.

## Notes

### Competing Interest Statement

All authors have completed the ICMJE uniform disclosure form. All authors declare the following competing interests: this work was completed by the authors while employed at, or while collaborating with, Meta.

### Author Declarations

An external review by Bluebonnet Ethical Review determined that this activity is appropriately classified as Not Human Subjects Research (NHSR) and that IRB review is not required.

### Summary of Updates

Supplemental eMethods updated with additional information. Additional author added for statistical analysis.

